# Performance Evaluation of the Elecsys HCV Duo Immunoassay in the Public Healthcare setting in Cape Town, South Africa

**DOI:** 10.1101/2025.03.14.25323950

**Authors:** Diana R Hardie, Stephen NJ Korsman, Ziyaad Valley-Omar, Nadia Petersen, Russell Cable, C. Wendy Spearman, Mark Sonderup

## Abstract

**Objectives:** Improved hepatitis C virus (HCV) diagnosis and linkage to care is crucial to achieve WHO 2030 elimination targets. Simplification of diagnostics remains key. We evaluated the performance of Elecsys HCV Duo antigen/antibody immunoassay in patients using public healthcare in Cape Town, South Africa.

**Methods:** 253 HCV seropositive and 214 seronegative samples were tested, and results correlated with standard-of-care (SOC) serology, RNA, viral genotype, patient demographics and disease markers. Thirteen patients on antiviral-therapy were also evaluated.

**Results:** Elecsys HCV Duo antibody was equivalent to SOC serology, while antigen had 100% negative percent agreement in non-viraemic samples. One incident infection with viral load of 54000 IU/mL was antigen positive/antibody negative. Overall, antigen detection was 62.2% in RNA-positive samples. Viral load strongly predicted reactivity, with antigen positive rates of 17.5% (<5 log IU/mL), 71.4% (5-6 log IU/mL), 89.4% (6-7 log IU/mL) and 100% (>7 log IU/mL). Antigen detection in genotype-1 infections was significantly better at 69.6% (95% CI 59.5-79.7) than for non-genotype-1 at 43.2% (95% CI 28.7-57.7). In treated patients, antigen mirrored RNA clearance but was only reliable if positive at baseline.

**Conclusion:** Elecsys HCV Duo detected active infection in 62% of viraemic patients and in 70% with genotype-1. In our cohort, 49% of newly diagnosed patients would require RNA testing.

**Highlights:**

- Elecsys HCV Duo detected active infection in 63% of new HCV RNA positive patients
- Genotype-1 status and RNA >6 log IU/mL are determinants of antigen positivity
- In antiviral therapy, antigen mirrors RNA decay, and is useful if baseline positive
- 49% of new HCV patients would still require RNA test to determine viraemia

## Introduction

The burden of disease from hepatitis C virus (HCV) infection is significant. Approximately 59 million people are persistently infected globally [1], 20% of whom are in Sub-Saharan Africa [2]. In South Africa (SA), prevalence is under-studied, but an estimated 600 000 people have chronic infection [3]. Risk factors, namely unsafe medical practices, re-use of needles, and unsafe sex, especially in men who have sex with men (MSM) who are HIV coinfected drive new infections in South Africa. In people who inject drugs (PWID), HIV coinfection is highly prevalent with shared modes of transmission [4].

Overall, less than 20% of patients know their status [4]. Patients are often from marginalised communities and don’t readily access healthcare services [4]. Improved diagnosis and rapid linkage to care is critical to achieve the WHO 2030 HCV elimination targets (90% infections diagnosed, 80% on therapy and 65% reduction in morbidity) [5]. Control of HCV should be achievable, with increasing availability globally of directly acting antiviral (DAA) therapy.

In South Africa, HCV infection is mostly diagnosed by centralised antibody testing, with confirmation of viraemia requiring a separate bleed for viral load (VL). VL is sensitive and necessary for blood and organ donor screening, but this approach is costly and results in significant loss to follow-up.

The WHO recommends core antigen testing as a surrogate where RNA testing is unafordable or unavailable [6]. The Abbott ARCHITECT HCV Ag (Abbott Park, Chicago, IL) quantitative assay has been well studied in this role, but is costly and impractical for routine screening, particularly in low prevalence settings like ours. Where prevalence is ≤5%, confirmatory VL is required for antigen positives [7].

The Roche Elecsys HCV Duo immunoassay is a 5^th^ generation electro-chemiluminescence immunoassay (ECLIA) which detects HCV core antigen and anti-HCV antibody simultaneously. It uses peptides and recombinant antigens derived from core, NS3 and NS4 proteins to capture anti-HCV antibody, and anti-core monoclonal antibodies to detect HCV antigen in blood [8]. This major diagnostic advance aims to fill two critical needs, namely to identify HCV infection and detect viraemia vs resolved infection in new patients. Our aim was to evaluate assay performance (diagnostic accuracy) in our setting, on routine samples, with various genotypes (GT) and to assess its utility for monitoring patients on DAA treatment.

## Methods

### Ethics approval

was obtained from the University of Cape Town Human Research Ethics Committee, HREC REF 828/2023.

#### Study design

HCV antibody positive routine diagnostic samples were prospectively collected at the NHLS diagnostic virology laboratory at Groote Schuur Hospital in Cape Town, South Africa. This laboratory serves the public healthcare sector, receiving samples from hospitals and clinics in the greater Cape Town area.

Residual samples submitted for HCV testing between June 2023 and March 2024 were archived and tested by Elecsys HCV Duo on the COBAS e801 at the Western Cape Blood Service in Cape Town. Manufacturer defined values and cut of indices (COI) were used to determine results. Data was collected on clinic location, patient demographics, presence of coinfections (HIV and HBV) and biochemical parameters.

Samples had been screened with Roche COBAS e601 HCV antibody as standard of care (SOC). Antibody positive samples were frozen at -20 °C in paired aliquots (for serology and molecular testing). HCV VL was tested, either as routine care on GeneXpert HCV (Cepheid, Sunnyvale, CA), or later, if data was not available. In this case a stored aliquot was tested by Roche HCV COBAS 5800.

HCV VL assay concordance was verified by testing 10 viraemic samples by both assays.

HCV genotyping was performed on a subset of RNA positive samples, either during clinical care, or for study purposes, to evaluate assay performance for diferent genotypes. In addition, 51 archived samples of known genotype (GT) and VL were included, to increase the proportion of non-GT1 samples. These were from patients diagnosed prior to study onset with available residual sample at -20 °C. The HCV genotyping methodology used was Sanger sequencing of part of the NS5B gene. Selected samples were serially diluted in HCV negative serum to determine antigen detection endpoints.

### Precision testing

A subset of antigen screen positives and samples with COI close to the cut-of, were retested in duplicate to evaluate assay precision.

### Patients on DAA therapy

Patients who were starting DAA therapy (between January 2024 and July 2024) were recruited from the Liver Clinic at Groote Schuur hospital and consent was obtained to collect samples for HCV Duo testing at baseline, 4 weeks, end of therapy (12 weeks) and 12 weeks after EOT, in addition to SOC VL testing. In total, 13 patients were enrolled.

### HCV seronegative samples

214 HCV antibody screen-negative samples from patients with acute hepatitis were collected over 2-months. Samples were sourced from sites of higher HCV prevalence and from patients between 18-40 years, as HCV prevalence in Cape Town is highest in this group.

### Statistical analysis

Statistical analysis was performed using Excel and Graphpad Prism. Descriptive statistics including median, interquartile ranges were used to define non-parametric variables such as age, viral load and patient biochemical and haematological parameters. 95% confidence intervals were used to define measurement uncertainty. Bland-Altman plot was used to compare the 2 viral load assays and receiver operator curve analysis to define the COI that gave best assay sensitivity and specificity.

## Results

### HCV positive diagnostic samples

During the study period, 10084 HCV antibody tests were performed in our laboratory and 229 patients with newly diagnosed HCV infection identified, giving an HCV seroprevalence of 2.3%.

Of these, 63 samples were insuficient for further testing, but with the additional stored samples, n=253 HCV antibody screen positive samples were available for evaluation. The median age of antibody positive patients was 37 [IQR 32-46] years, with 183 (72.9%) male (**Figure 1**). Of note, 41% of patients were coinfected with HIV and 8.5% with hepatitis B.

**Figure 1:**
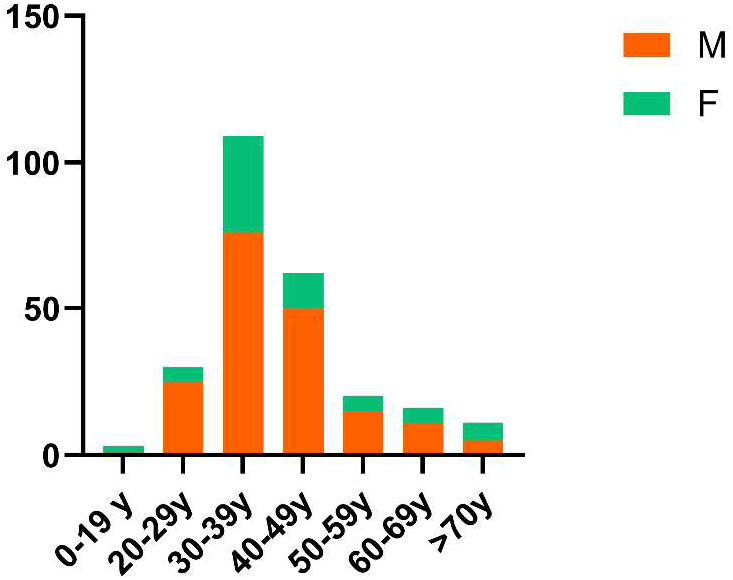
Age and sex distribution of 251 confirmed HCV antibody positive patients whose samples were used for this study. Median age [IQR] for males is 38 [32-45] and for females 37 [33-48].

Baseline patient characteristics are seen in **Table 1**.

**Table 1:** Patient characteristics.

|  | HCV antibody positive patients<br>n= 251# | HCV antibody negative patients<br>n=214 | DAA patients (baseline)<br>n=13 |
| --- | --- | --- | --- |
| Sex: |  |  |  |
| Male (%) | 183(72.9) | 103 (48.1) | 10 (76.9) |
| Female (%) | 68 (27.1) | 111 (51.9) | 3 (23.1) |
| Age (median [IQR]) | 37[32-45] | 31 [26-36] | 46 [38-55] |
| RNA status: |  | ND |  |
| RNA negative (%) | 50 (19.9) |  | 0 |
| RNA positive (%) | 201 (80.0) | 1 | 13 (100) |
| Viral load (median [IQR]) | 5.66 [4.79-6.33] |  | 6.02 [5.76-6.59] |
| Genotype known | 123 (61.2)* | 1 | 13 (100) |
| Genotype 1 (%) | 79 (64.2)^ | 1 | 8 (62) |
| Other genotype | 44(35.8) |  | 5 (38) |
| HIV status: |  |  |  |
| Positive (%) | 103 (41.2) | 65 (30.4) | 4(30.7) |
| Median CD4 count | 305 [169-438] | 75 [26-235] | 650 |
| Negative (%) | 118 (47.2) | 120 (56.1) | 9(69.3) |
| Unknown (%) | 28 (11.2%) | 30 (14.0) | 0 |
| HBV status: |  |  |  |
| Positive (%) | 21 (8.4) | 12 (5.6) | 0 |
| Negative (%) | 205 (82) | 197 (92.0) | 13 (100) |
| Unknown | 24 (9.6) | 7 (3.3) | 0 |
| Creatinine $\mu\text{mol/mL}$ [IQR] | 60 (54-86) | 68 [53-101] | 82 [57-118] |
| ALT U/L [IQR] | 36 [19-65] | 40 [20-211] | 67 [35-116] |
| AST U/L [IQR] | 48 [28-77] | 68 [31-303] | 66 [27-150] |
| TBR $\mu\text{mol/mL}$ [IQR] | 10 [6-38] | 16 [7-50] | 8 [7-18] |
| Albumin g/L [IQR] | 30 [21-38] | 30 [24-39] | 42 [36-45] |
| WCC $\times 10^9/\text{L}$ [IQR] | 10.29 [7.2-16.3] | 8.14 [5.77-11.6] | 6.38 [5.1-8.2] |
| Haemoglobin g/dL [IQR] | 11.0 [9.1-12.7] | 11.6 [9.3-13.3] | 15.2 [13.0-16.3] |
| Platelets $\times 10^9/\text{L}$ [IQR] | 267 [165-379] | 286 [212-362] | 248 [155-301] |

\* of RNA positive samples
^ of genotyped samples

### HCV viral load in study samples

HCV VL had been tested by GeneXpert on 166 samples and 87 were tested by Roche HCV COBAS 5800. Based on a verification the assays were judged to be equivalent (a slight positive bias of 0.118 with HCV COBAS 5800) as shown in the Bland Altman plot, **Figure 1S**. Overall, 20.5% (52) of patients who were anti-HCV antibody positive had undetectable VL (no active infection). In the 201 viraemic patients, VL ranged from 1.74 to >7 log IU/mL. The median VL in RNA positive samples was 5.67 [IQR 4.85-6.34] log IU/mL.

### HCV Duo antibody performance

251 of 253 HCV screen positives were antibody positive on Elecsys HCV Duo, giving a positive percent agreement (PPA) of 99.2%. Two discordant samples with low screening COI values which were HCV RNA negative, were judged to be screen false positives. 50 (19.9%) patients were judged as having previous HCV infection.

All 214 screen HCV antibody negative samples were negative on Elecsys HCV Duo antibody, giving a negative percent agreement (NPA) of 100%.

### Elecsys HCV Duo antigen performance

Antigen was negative on all 50 confirmed HCV antibody positive samples with undetectable HCV VL, giving a NPA of 100%.

In RNA positive samples, antigen positivity varied according to VL. The median HCV VL in antigen positive samples was 6.14 [IQR 5.55-6.56] log IU/mL while that of negative samples was lower, at 4.54 [IQR 3.39-5.34] log IU/mL, p<0.0001 **(Figure 2a)**.

**Figure 2:**
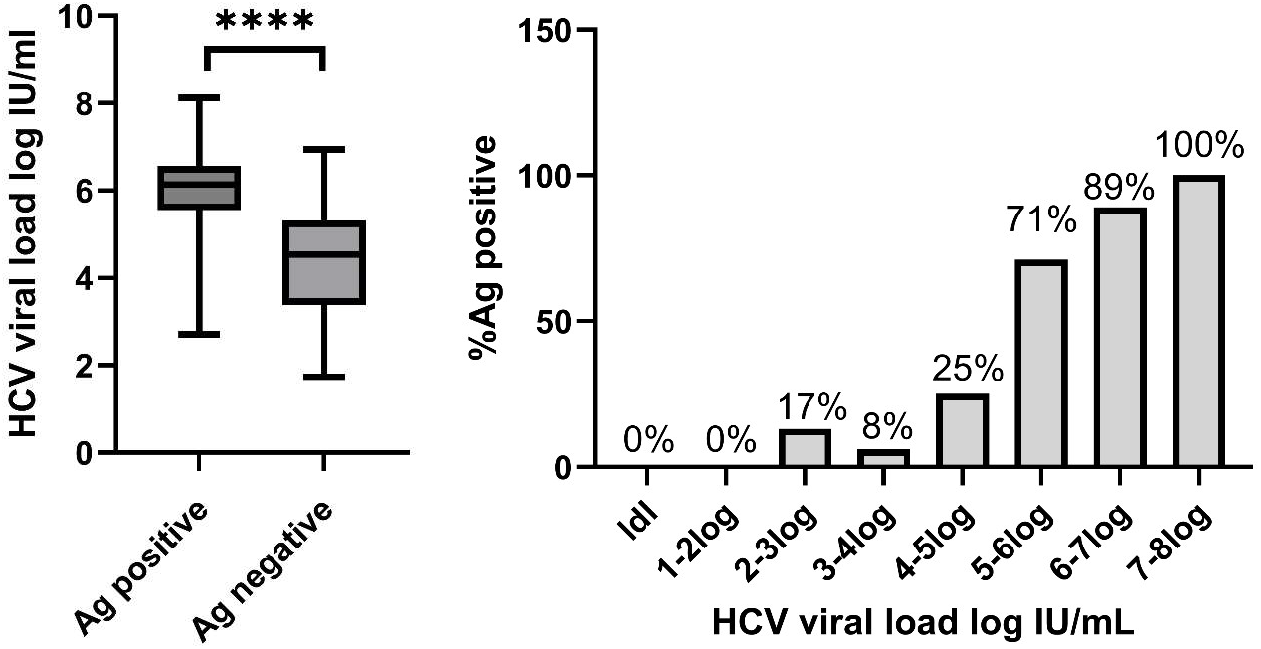
HCV viral load in Elecsys HCV Duo antigen positive and negative HCV positive diagnostic samples. In (a) the median viral load in antigen positive (6.14 [IQR 5.55-6.56]log IU/mL) and negative (4.54 [IQR 3.39-5.34] log IU/mL) samples. Whiskers reflect the maximum and minimum viral load range. Ag= antigen. In (b) antigen percentage positivity by viral load category in HCV antibody positive diagnostic samples. Antigen positivity rate increased from 17.5% (<5 log IU/ml) to 71.4% (5-6 log IU/mL) to 89.4% (6-7 log IU/mL) to 100% (>7 log IU/mL).

Overall, sensitivity of Elecsys HCV Duo for identifying active infection in the 201 patients with detectable HCV RNA (125/201) was 62.2% (95% CI 56.6-70.1). (**Table 2**) Antigen detection was highly dependent on VL. Where VL was below 5 log IU/mL, antigen was detected in only 17.7% (11/62) but improved to 71.4% (45/63) for samples with VL of 5-6 log IU/mL and 89.4% (59/66) with VL of 6-7 log IU/mL. All 10 samples with VL >7 log IU/mL were detected. (**Figure 2b)**

**Table 2.** Contingency table of HCV Duo antigen detection by HCV RNA status.

|  |  | HCV RNA status |  |  |
| --- | --- | --- | --- | --- |
|  |  | Positive | Negative | Total |
| Elecsys HCV Duo antigen | Positive | 125 | 0 | 125 |
|  | Negative | 76 | 50 | 126 |
|  | Total | 201 | 50 | 251 |

### Elecsys HCV Duo antigen performance according to HCV genotype

HCV GT and sub genotype was known on 123 (61.2%) of 201 RNA positive samples. N=79 were GT1 (75=1a and 4=1b) and 44 were other GTs. The most frequently detected other GTs were 3a (18), 4 (10), 5a (13) and 2 (3). Antigen detection in patients with GT1 infection was 69.6% (95% CI 59.5-79.7). This was significantly better than in patients with non-GT1 where antigen was detected in only 43.2% (19/44) of samples (95% CI 28.7-57.7), p=0.0068. By GT, antigen was positive in 4/18 (22%) patients with GT3a, 6 of 10 (60%) with GT4, 8 of 13 (62%) with GT5a and 1 of 3 (33%) with GT2. **Figure 3** plots antigen detection status by VL for GT1 and non-GT1 samples. Non-GT1 samples are colour coded for clarity.

**Figure 3:**
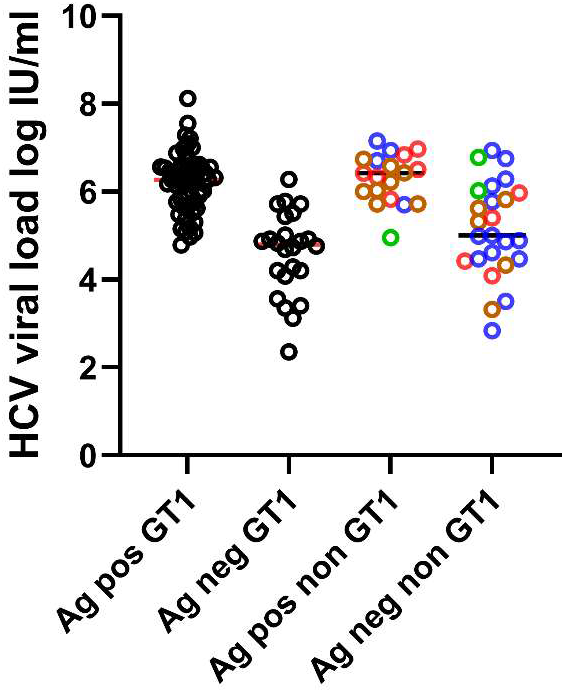
Antigen status and viral genotype: Scatter plot shows antigen detection by viral load in GT1 and non-GT1 clinical samples. Median viral load in positive and negative samples is similar in both GT1 and non-GT1 samples. The GT of non-GT1 samples is indicated by colour coding where green indicates GT2, blue GT3a, red GT4 and brown GT5a.

The median VL in antigen positive GT1 samples was 6.27 and 4.80 log IU/mL in negative samples. For non-GT1 samples, the median VL in positive samples was 6.42 and 5.00 log IU/mL in negative samples. This suggests that reduced detection of non-GT1 infections was not primarily caused by VL diference.

### Antigen cutof index (COI)

The antigen assay gives a qualitative readout, with COI ≥1 indicating a positive result. In positive samples, values varied from 1 to 211. Some RNA positive samples had COI values in the negative range close to the cutof, (Figure 2S) suggesting that sensitivity could be improved by adjusting the COI or creating a grey zone. This was evaluated by receiver operator curve (ROC) analysis. ROC analysis gave an area under the curve (AUC) of 0.798 (95% CI 0.7459 to 0.8501). **Figure 3S**. Based on our data, use of a COI of 0.85 gave the best discrimination, with a PPA of 63.7% and NPA of 100%.

### Precision testing of antigen COI

Precision testing was performed on 51 samples with initial screen COI values in the high negative to positive range. All 15 samples with values around the cut-of on initial screening (0.8-1.5) were retested in duplicate, as were 35 samples with COI >1.5. Precision was good in samples with COI > 1.5 but, substantial variability was observed around the cut-of. Of the 15 samples in this range, 7 changed from positive to negative or negative to positive on repeat. All 15 were RNA positive. COI values are shown in **Table 3**.

**Table 3.** Antigen COI of samples where status changed on repeat testing.

| Screen COI | Repeat 1 COI | Repeat 2 COI | HCV VL log IU/mL |
| --- | --- | --- | --- |
| 0.863 | 1.70 | ins | 5.51 |
| 1.09 | 0.72 | ins | 4.63 |
| 0.962 | 1.15 | 1.13 | 5.72 |
| 1.11 | 0.793 | 0.837 | 5.66 |
| 0.805 | 0.952 | 0.993 | 4.76 |
| 1.28 | 1.00 | 0.992 | 5.83 |
| 1.04 | 0.80 | 0.782 | 5.87 |
| 1.30 | 0.896 | 0.818 | 6.98 |
| 1.49 | 1.02 | 0.969 | 4.95 |

### Relationship between antigen COI and viral load

To evaluate the relationship between antigen COI and VL, log transformed VL and COI values were plotted. There was a significant positive correlation, with the slope of the regression line of 0.531 (95% CI 0.302-0.760). **Figure 4S**

### Antigen COI in serially diluted samples

Limit of detection was also evaluated in serially diluted samples. 10 samples (of GTs 1a, 1b, 4c, 4q, 4r and 5a) were diluted ten-fold in negative human serum. The mean VL at the detection endpoint was 5.6 log IU/mL (SD 0.56). In contrast, antibody remained detectable at 1:10^−3^ to 1:10^−4^. As for undiluted samples, there was a modest correlation between antigen COI and HCV VL (slope 0.7096). **Figure 5S**

### Antigen kinetics in DAA patients

Of the 13 DAA patients, median age was 46 [IQR 41-51] years, with median baseline HCV VL of 6.02 [IQR 5.76-6.59] log IU/mL. 10/13 were males and 4/13 were HIV coinfected. In terms of genotype, n=6 were 1a, n=2 1b, n=1 2c, n=3 were 3a and n=1 was 5a. All received12 weeks of daily fixed dose sofosbuvir and velpatasvir. All responded to therapy, with sustained virological suppression (SVR). Typical VL kinetics were observed, with a 4-5 log decay at week 4, and undetectable VL at week 12 (EOT).

Antigen was positive at baseline in 7/13 patients (negative in patients with GTs 1a, 3a, 2c and 5a). In 4/7 baseline positive patients, antigen was negative at week 4. In 3 patients, antigen remained detectable at week 4, despite low VLs of 48, 72 and 208 IU/mL respectively.

(**Table 1S** shows antigen COI and VL data of 7 baseline positive patients during treatment.)

### Seronegative patients screened for incident HCV

214 samples from HCV seronegative individuals were evaluated. 213 were both antibody and antigen negative on the Elecsys HCV Duo. One sample was positive only for antigen, COI=2.18, with antibody COI= 0.442 and HCV VL of 5.73 log IU/mL. Thus, in this cohort, only one incident infection was identified.

## Discussion

Data is limited on HCV prevalence in patients who access public healthcare in South Africa. We observed a prevalence of 2.2% over the 10-month study period. Most HCV positive patients were young (median age 37), from impoverished communities, in poor health (based on laboratory data), with a high likelihood of comorbidities (41% with HIV and 8.5% with HBV). Seventy-two percent were male. These features fit the recognised HCV risk profile globally [1]. Our screening algorithm involves centralised serology with non-reflexive follow-up VL on positive patients [9]. Using this approach, many patients are lost to follow up. Indeed, during the study, VL was only routinely performed on 56.6% of newly diagnosed patients. Dual antigen/antibody testing holds great promise, if it can reliably identify viraemia during primary screening. We evaluated the Elecsys HCV Duo 5^th^ generation assay and found that the antibody showed good equivalence with the SOC assay, with a PPA of 99.1% and 100% NPA in screen negative samples. Similarly, core antigen had a 100% NPA in non-viraemic samples, both RNA negative seropositive and seronegative. The assay detected one incident infection in an HCV antibody negative sample with VL of 54000 IU/mL.

Overall, antigen confirmed viraemia in 62.2% of 201 RNA positive samples. Antigen positivity was strongly influenced by VL, increasing from 17.7% where VL was below 5 log IU/mL, to 100% where VL was >7 log IU/mL. Sensitivity was lower than in other recent studies, where detection rates between 70-73% of viraemic samples are reported. A lower average HCV VL in our samples, only 5.67 log IU/mL, could be responsible. The median VL in samples in the study by Bui et al. [10] was 6.3 log IU/mL and 6.5-7.0 log IU/mL in the study by Kanokudom et al [11]. The reason for the lower average viraemia in our patients is not clear. They were reflective of the population that we normally screen. Based on this data, viraemia would have been confirmed in 51% of these patients with the Elecsys HCV Duo, while 49% would have been antigen negative and required a VL.

Though not quantitative, the antigen COI showed modest correlation with VL. Precision was good, with greatest variability around the cut-of. Most samples with COI in the high negative range were RNA positive and ROC analysis of our data, showed an adjusted COI of 0.85 could improve sensitivity without sacrificing specificity. We suggest use of a grey zone to improve sensitivity.

While the assay detected viraemia of a range of GTs (1a and b, 3a, 4 and 5a), GTs other than 1 were detected at lower frequency. When only samples of known GT were evaluated, assay sensitivity in GT1 was 69.6% and only 41% in non-GT1 samples. As the mean VL in both GT1 and non-GT1 groups were equivalent, a diference in VL did not explain this. We did not have enough samples to examine each GT individually, but GT 3a was least likely to be detected. Lower detection of this GT was also noted with the Abbott Architect core assay [12] and also with Elecsys HCV Duo in the Kanokudom study (where the sensitivity for GT3a, at 62% was the lowest of all GTs) [11]. In another study, using the Abbott Architect antigen assay, lower antigen detection of GT3 and 6 samples was linked to an amino acid substitution (threonine to alanine/valine) at position 49 in the viral core protein [13]. The authors speculated that this substitution could result in reduced binding to capture antibodies in the test.

Lastly, we evaluated antigen kinetics in patients on therapy. In the 7 baseline positive patients, antigen mirrored VL but with slower clearance at week four in some. This likely reflects residual free core protein in subviral particles or immune complexes [14]. All patients had an SVR, with antigen correctly confirming EOT results. However, antigen testing cannot be used in for this purpose when negative at baseline.

### Study limitations

Testing was not performed in real-time thus it was not possible to evaluate the clinical impact of having results available to clinicians. Further study where the assay is used for primary screening should address this. GT data was only available for a subset of samples which reduced our ability to evaluate samples of GTs other than 1 individually. Due to the prospective study design, only 13 patients on DAA therapy could be evaluated.

## Conclusion

The Elecsys HCV Duo immunoassay identified active infection in 63% of viraemic patients and 70% of those with GT1. In our cohort, 49% of newly diagnosed patients would still require follow up VL testing. Adjustment of the cut-of or setting of a grey zone that covers the high negative range could improve assay sensitivity.

## Ethical Approval statement

Ethics approval was obtained from the University of Cape Town Human Research Ethics Committee, HREC REF 828/2023 on 10 November 2023. This work was carried out in accordance with Declaration of Helsinki for experiments involving humans. Written informed consent was obtained from the 13 patients on DAA therapy who provided samples for Elecsys Duo testing at routine visits. None of them were minors (age range 35-75 years).

## Data availability

Data will be made available via an excel spreadsheet as a supplementary file

## Acknowledgements

We would like to thank Mr Tyronne Jensen (Western Cape Blood Service) for technical assistance with HCV Duo testing and Ms Tathym Gelderbloem (National Health Laboratory Service) for assistance with HCV VL testing. Diagnostic kits to perform HCV Duo testing and HCV VL testing on a proportion of samples were provided by Roche Diagnostics.

