## supplementary data for "Performance Evaluation of the Elecsys HCV Duo Immunoassay in the Public Healthcare setting in Cape Town, South Africa"

**Supplementary files:**

**Figure 1S: Bland Altman plot of GeneXpert vs Roche cobas 5800 HCV viral load**

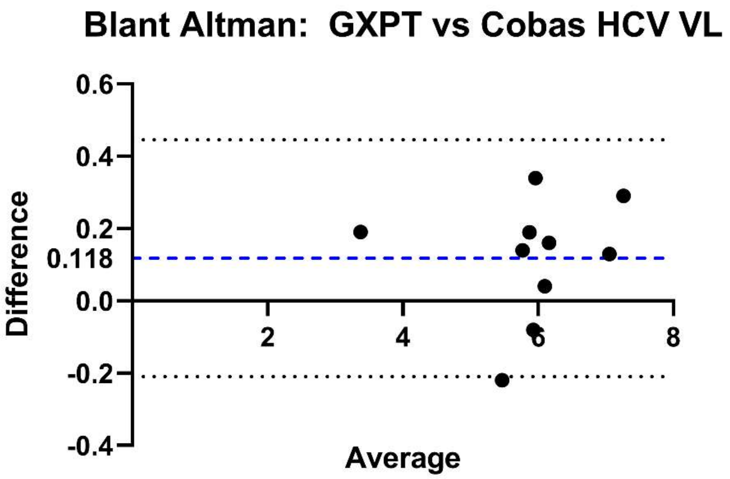


| | |
|---|---|
| Bias | 0,1180 |
| SD of bias | 0,1671 |
| 95% Limits of Agreement | |
| From | -0,2094 |
| To | 0,4454 |

**Table 1S**

**Antigen COI values over the course of treatment in baseline positive DAA patients**

| Patient | GT | Visit | Viral load | | Antigen COI |
|---|---|---|---|---|---|
| DAA01 | 1b | baseline | 1257854 | 6,10 | 7,29 |
| | | week 4 | LDL | 0,00 | 0,481 |
| | | EOT | LDL | 0,00 | 0,537 |
| | | 12 weeks post EOT | LDL | 0,00 | NT |
| DAA02 | 1a | baseline | 1894193 | 6,28 | 7,93 |
| | | week 4 | 72 | 1,90 | 1,3 |

| | | | | | |
|---|---|---|---|---|---|
| | | EOT | <10 | 1,00 | 0,721 |
| | | 12 weeks post EOT | LDL | 0,00 | NT |
| DAA03 | 1a | baseline | 174718 | 5,24 | 1,16 |
| | | week 4 | LDL | 0,00 | 0,528 |
| | | EOT | LDL | 0,00 | 0,515 |
| | | 12 weeks post EOT | LDL | 0,00 | NT |
| DAA04 | 1a | baseline | 9971367 | 7,00 | 2,27 |
| | | week 4 | 207 | 2,32 | 3,08 |
| | | EOT | LDL | 0,00 | 0,489 |
| | | 12 weeks post EOT | NT | | NT |
| DAA05 | 1a | baseline | 334000 | 5,52 | 6,59 |
| | | week 4 | 16 | 1,20 | 0,573 |
| | | EOT | LDL | 0,00 | 0,538 |
| | | 12 weeks post EOT | LDL | 0,00 | NT |
| DAA06 | 1a | baseline | 10083519 | 7,00 | 46,9 |
| | | week 4 | 117 | 2,07 | 0,522 |
| | | EOT | 10 | 1,00 | 0,532 |
| | | 12 weeks post EOT | LDL | 0,00 | NT |
| DAA07 | 1b | baseline | 3870000 | 6,59 | 151 |
| | | week 4 | 48 | 1,68 | 6,51 |
| | | EOT | LDL | 0,00 | 0,573 |
| | | 12 weeks post EOT | NT | #VALUE! | NT |

**Figure 2S**

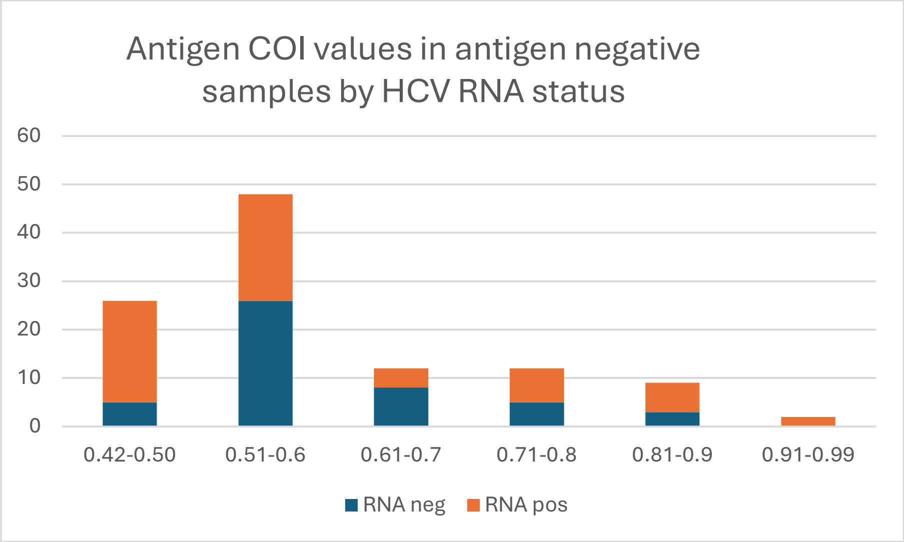


**Figure 2S: Shows COI ranges for antigen negative HCV antibody positive samples stratified by RNA status. In the high negative range, >0.7 the proportion of RNA positive samples rises.**

**Figure 3S**

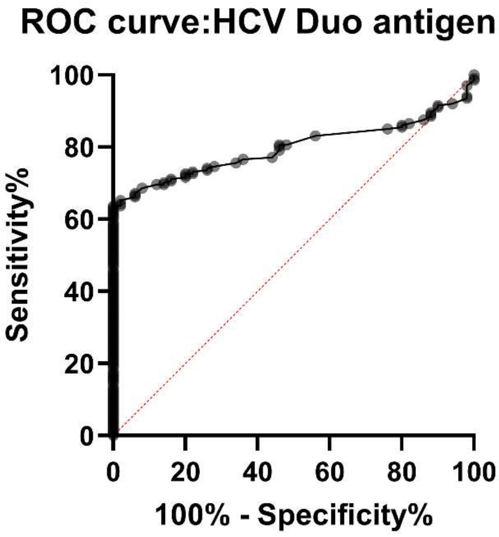


Area under the ROC curve

| | |
|---|---|
| Area | 0,7980 |
| Std. Error | 0,02659 |

| | |
|---|---|
| 95% confidence interval | 0,7459 to 0,8501 |
| P value | <0,0001 |
| Data | |
| Controls (RNA neg OD) | 50 |
| Patients (RNA pos OD) | 201 |

**Figure 4S**

**Relationship between COI and HCV viral load in positive samples**

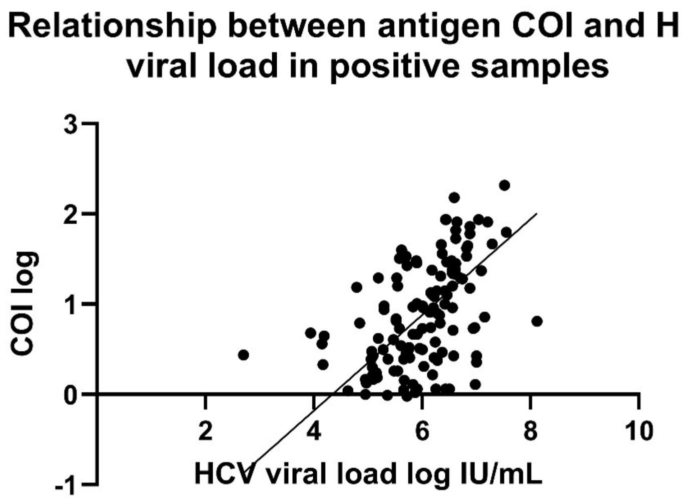


**Figure 4S: Log COI and HCV values plotted using Deeming regression analysis shows a slope of 0.531 with intercept at -2.304. Y = 0,5310*X - 2,304**

**Figure 5S Antigen signal in sample dilutions**

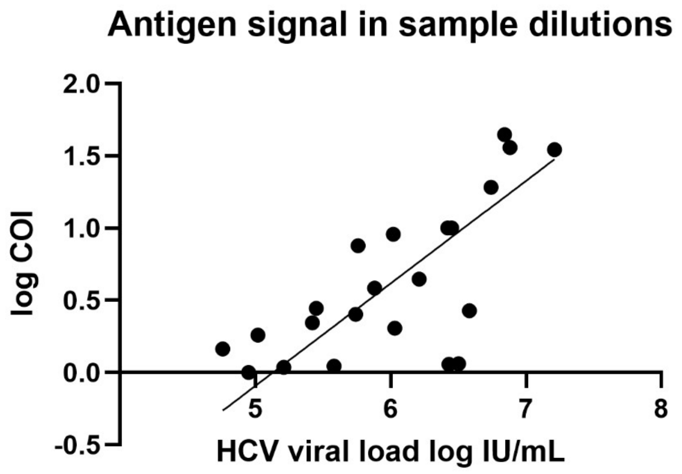

**Figure 5S: Log COI and HCV viral load values of diluted samples are plotted. show correlation with slope of 0.710, (95%CI 0.4502-0.9689). Y = 0.7096*X - 3.639**
